# Association between Accelerometer-Measured Irregular Sleep Duration and Longitudinal Changes in Body Mass Index in Older Adults

**DOI:** 10.1101/2024.10.30.24316315

**Authors:** Sina Kianersi, Kaitlin S. Potts, Heming Wang, Tamar Sofer, Raymond Noordam, Martin K Rutter, Susan Redline, Tianyi Huang

## Abstract

Irregular sleep duration may disrupt circadian rhythms and contribute to metabolic, behavioral, and mood changes, potentially increasing the risk for obesity. However, quantitative data on the relationship between sleep duration irregularity and weight change are lacking. In this prospective study, we analyzed data from 10,572 participants (mean age: 63 years) in the UK Biobank who wore accelerometers for a week between 2013–2015 and had two body mass index (BMI; kg/m²) measurements on average 2.5 years apart. Irregular sleep duration was assessed by the within-person standard deviation (SD) of 7-night accelerometer-measured sleep duration. Participants with sleep duration SD >60 minutes versus ≤30 minutes had 0.24 kg/m^2^ (95% CI: 0.08, 0.40) higher BMI change (kg/m^2^), standardized to three-year intervals, and 80% (95% CI: 1.28, 2.52) higher risk for incident obesity, after adjusting for sociodemographic factors, shift work, and baseline BMI or follow-up period (*p*-trend<0.02 for both). These associations remained consistent after adjusting for lifestyle, comorbidities, and other sleep factors, including sleep duration. Age, sex, baseline BMI, and genetic predisposition to higher BMI (measured with a polygenic risk score) did not appear to modify the association. Since irregular sleep duration is common, trials of interventions targeting sleep irregularity might lead to new public health strategies that tackle obesity.

## INTRODUCTION

The World Health Organization estimated that in 2016 at least 650 million adults worldwide, corresponding to 13% of the global adult population, were classified as obese, defined as a body mass index (BMI) of at least 30 kg/m^2^ (1). Obesity prevalence has almost tripled since 1975 (1), with projections suggesting that over one billion individuals will be affected by 2030 (2). Increased BMI is causally associated with a multitude of adverse health outcomes, including cardiovascular diseases, diabetes, certain types of cancer, and premature death (3, 4). Lifestyle modification represents a cornerstone for both the prevention and initial clinical management of obesity (5), but is mainly focused on nutrition and physical activity. Identification of novel lifestyle factors contributing to obesity could enhance public health and clinical strategies to reduce the burden of this condition.

Sleep plays a key role in daily energy metabolism with short sleep duration as an established risk factor for obesity (6-8). However, less is known about other dimensions of habitual sleep, particularly sleep regularity, an emerging component of healthy sleep (9). Irregular sleep, marked by daily fluctuations in the duration or timing of sleep, is increasingly recognized as a distinct risk factor for metabolic health, independent from sleep duration or poor sleep quality (9). Sleep irregularity is likely a consequence of multiple factors, including modern working patterns, recreational activities (e.g., screen time), sleep-disrupting substances (e.g., caffeine, alcohol, and nicotine), social interactions, environmental disturbances, and emotional stressors (e.g., depression and anxiety) (6). In the general population, chronically irregular sleep patterns may disrupt circadian rhythms, causing circadian misalignment and leading to long term adverse health consequences. Circadian misalignment (i.e., misaligned feeding-fasting or sleep-wake cycles) challenges metabolic health, as it could dysregulate appetite hormone secretion, reduce energy expenditure, change energy intake timing, and foster poor dietary choices (6). In recent years, a growing body of literature has shown associations between irregular sleep and cardiometabolic disorders (10), including type 2 diabetes mellitus (11), and cardiovascular diseases (12). Besides, genetic variants, particularly when coupled with certain environmental factors, considerably contribute to the development of obesity (13, 14). Previous studies have identified sleep duration as an environmental modifier, with both short and long sleep durations exacerbating genetic influences on obesity (15). Similarly, a genetic predisposition to obesity may influence the relationship between irregular sleep patterns and weight changes. However, the dynamics of this interaction remain largely unexplored.

Irregular sleep has been implied as a potential risk factor for obesity (9). Systematic reviews, however, have identified methodological limitations in the literature, including variation in methodology for quantifying regularity (with the intra-individual standard deviation (SD) metric offering the advantage of being particularly easy to interpret (16)), small samples, and mostly cross-sectional study designs, which provide limited understanding of temporal relationships (9, 17, 18). Cross-sectional studies, including one particularly large study involving 120,522 adults, found a positive association between sleep duration variability and prevalent obesity (19-22). Three longitudinal studies to date, one of which used self-reported data to assess irregular sleep durations, also suggested that higher variations in sleep duration is associated with weight gain and increased obesity risk (23-25). Despite these initial studies supporting a role of sleep irregularity in obesity, one of the systematic reviews using the GRADE framework, a systematic approach for evaluating the quality of evidence (26), assessed the quality of included studies as ranging from low to very low (17), indicating the necessity for a comprehensive longitudinal study utilizing objectively measured variables.

In this study, we investigated the longitudinal association between accelerometer-measured sleep duration regularity and BMI change as well as incident obesity in a large sample of UK adults. Based on the above-mentioned literature, we hypothesized that individuals with more irregular sleep duration would experience greater BMI increase and a higher risk of incident obesity over time, than those with more regular sleep duration. Further, we explored how genetic predisposition to obesity influences the association between irregular sleep and incident obesity.

## METHODS

### Study design, setting, and participants

The UK Biobank (UKB) is a prospective cohort study that recruited participants aged 40-69 years between 2006 and 2010 (27). As of July 2023 (data version 16.1) UKB includes 502,364 participants. This cohort was drawn from 9.2 million people registered with the National Health Service (NHS), including a wide range of sociodemographic backgrounds from across the UK. Every participant consented electronically and underwent a baseline assessment at one of 22 centers nationwide. This assessment entailed a computer-assisted interview covering sociodemographic, psychosocial, and lifestyle factors, a touch-screen questionnaire, along with physical evaluations and the collection of biological samples including blood, urine, and saliva (27). For a small subset of the cohort, these baseline evaluations were repeated up to three additional times between 2012-2024. Baseline assessment also included genotyping and the conduct of standard biochemical tests. Ongoing health outcomes are tracked through linkages to national health databases, including Hospital Episode Statistics, primary care, and death registry records. The UKB received approval from the National Health Service (NHS) North-West Multi-Centre Research Ethics Committee.

From 2013-2015, a total of 236,519 UKB participants who had valid email addresses were invited to join an accelerometer study (28). The response rate was 44% and 103,628 participants provided accelerometer data. Among 92,096 participants with good quality sleep data derived from accelerometers (e.g., well calibrated raw data), 11,575 with two BMI measurements (see BMI assessment below for details) were included in the current study (Supplemental Figure 1). Additionally, we excluded participants with <5 nights of sleep duration data (n = 312), sleep duration SD >4 hours (n = 2), extreme BMI changes based on the generalized Extreme Studentized Deviate (ESD) method (n = 158) (29), or those whose two BMI measurements were less than 182 days (∼six months) apart (n = 531), leaving 10,572 participants for analysis on continuous BMI change outcomes. For analyses involving incident obesity, we excluded participants with a baseline BMI ≥30 kg/m^2^, leaving 7,937 participants for these analyses. Included and excluded participants had similar characteristics (Supplemental Table 1)

### Sleep duration standard deviation (exposure)

Participants in the accelerometer study were equipped with the wrist-worn waterproof triaxial Axivity AX3 accelerometer (Open Lab, Newcastle University) (28, 30). This device is known for its consistent performance in terms of inter- and intra-device variability in movement measurement and has shown to outperform the GENEActiv accelerometer, another device frequently utilized in health studies, during multi-axis shaking tests (28, 31). Following methodologies established in previous UKB research (32, 33), the processing of accelerometer data was conducted using the R package GGIR version 1.5-12, which was used to calculate daily sleep duration, defined as the total duration of sleep episodes within the main sleep period excluding naps (34, 35). A validation study involving 50 participants demonstrated that the GGIR package’s C-statistic for accurately identifying sleep period time windows based on accelerometer data compared to polysomnography data exceeded 0.8 (36). Participants had an average of 6.8 nights of derived sleep duration data (minimum = 5, Q1 = Q3 = 7 nights). The within-person standard deviation (SD) of sleep duration (hours) measured by the accelerometer across the nights worn was used to define irregular sleep duration. The sleep duration SD was modeled both as a continuous variable and in categories of ≤30, 31-45, 46-60, and >60 minutes. These categories were selected to improve the interpretability of the results, account for the data distribution, and align with recommendations from prior research (9).

### BMI assessment

During initial assessment center visits, healthcare technicians and certified nurses measured participants’ standing height and weight. Standing height was measured using a Seca 202 stadiometer (Hamburg, Germany), and weight was determined through a Tanita BC418MA body composition analyzer (Tokyo, Japan) for participants undergoing bioelectrical impedance analysis. For those who did not undergo this specific analysis, a standard scale was used for weight measurement (37). No dietary restrictions were enforced prior to weighing, and measurements were taken at various times throughout the day. BMI (kg/m^2^) was constructed using height and weight values. While nearly all UKB participants have baseline BMI measurements, our study utilized BMI measurements from follow-up assessment center visits (2012-2024) available to a smaller number of participants given the timeframe of the accelerometer study. Additionally, at the time of this study, approximately 45% of UKB participants had their primary care records, recorded by healthcare professionals in general practices, linked to their UKB data (38). Using data from assessment center visits and primary care records, we identified two BMI measurements for each participant. The baseline measurement was selected to be proximate to the accelerometer study (±1 year). The follow-up BMI measurement was chosen to be at least six months subsequent to the accelerometer study and the baseline BMI measurement. If multiple BMI measurements met these criteria, we chose the one closer to the accelerometer study for baseline measurement or the one closest to three years post-baseline for follow-up measurement (Supplemental Text).

Assuming that BMI changed at a constant rate and in a linear fashion during the follow-up period, we created three outcomes using the two BMI measurements. 1) BMI change (kg/m^2^), standardized to three-year intervals, was determined by dividing the difference between follow-up and baseline BMI measurements by the number of days between these two measurements and multiplying by 1095 days. 2) Relative BMI change percentage was defined by dividing the three-year standardized BMI change by the baseline BMI value, and then multiplied by 100 to convert it to a percentage (See the Supplemental Text for the relevant equations). 3) Incident obesity was defined as the transition of an individual’s BMI from below 30 kg/m^2^ at baseline to 30 kg/m^2^ or above at follow-up.

### Covariates

Sociodemographic factors, comprising age, ethnic background, sex, education, Townsend deprivation index, and occupation/shift work, were assessed at the UKB baseline assessment center visits from 2006-2010. Similarly, smoking, alcohol use, and certain sleep-related covariates (i.e., insomnia symptoms, chronotype, and daytime sleepiness) were self-reported at baseline, and the healthy diet score was computed from diet information collected at baseline using the UKB touch-screen questionnaire (39). A limited number of participants underwent repeated assessments during visits between 2012-2024. When available, we incorporated the closest covariate measurement to the accelerometer study date, considering data collected either before or within one year after this date. However, for most (N = 9,117), the information on covariates was acquired from their initial assessment center visit. The distributions of covariates were similar between participants with updated measurements and those without (Supplemental Table 3). Using accelerometers, physical activity levels and sleep duration were derived, with activity levels determined by the daily average of moderate-to-vigorous physical activity (minutes/day) (40). Sleep apnea cases were identified using ICD-10 code G47.3 (41). The following comorbid conditions diagnosed prior to the accelerometer study were included, as identified through hospital admission and/or primary care data (ICD-10 codes), or self-reports: dyslipidemia, hypertension, self-reported depression (based on PHQ-2 screening or a doctor visit for depression), and type 2 diabetes (41). Genotyping was conducted for UKB participants (42), and a polygenic risk score (PRS) for BMI was created by a previous study and returned to UKB. The PRS was trained from genome-wide summary statistics of a meta-analysis of independent genome-wide association studies using a Bayesian based approach (43, 44). A higher PRS for BMI suggests a greater genetic predisposition towards an increased BMI. Using tertile cutoffs for the PRS, we grouped participants into three categories: low BMI PRS (first tertile), intermediate BMI PRS (second tertile), and high BMI PRS (third tertile) (Supplemental Table 2 summarizes UKB data-fields implemented in our study).

### Statistical analysis

We reported the distributions of covariates across sleep duration SD categories using mean (SD) and frequency percentage. Multivariable-adjusted linear regression analyses were used to assess the associations between sleep duration SD and the continuous BMI change outcomes, estimating coefficients that quantified BMI change associated with both categorical sleep duration SD and each one-hour increase in sleep duration SD. We conducted the analysis separately for 3-year standardized BMI change and relative BMI percentage change, which both demonstrated distributions that approximated normality (Supplemental Figure 3). We fitted three multivariable-adjusted regression models. In model 1, we adjusted for age in years, sex, ethnic background, Townsend deprivation index, education, occupation/shift work, and baseline BMI. Given that metabolic comorbidities, depression, and behaviors could result from sleep duration SD, we analyzed these covariates in distinct models (2 and 3) to highlight possible over-adjustment for these models. In model 2, we further included smoking, physical activity, healthy diet score, alcohol consumption, dyslipidemia, hypertension, self-reported depression, and type 2 diabetes mellitus. In model 3, we added sleep apnea, insomnia, average sleep duration, chronotype, and daytime sleepiness, to account for other aspects of sleep. In all models, we designated the reference group as those with a sleep duration SD of 30 minutes or less, representing the group with the most regular sleep duration.

We fitted Poisson regression with a robust error variance to estimate the relative risk (RR) for the association between sleep duration SD and incident obesity. Here, we fitted the same three models, with additional adjustment of the time interval between the two BMI measurements as a covariate in all models to account for the fact that the follow-up period for incident obesity was not the same for all participants. We conducted two sensitivity analyses: 1) to understand the potential impact of unmeasured or unknown confounders on the observed associations, we estimated E values for analyses on incident obesity (45, 46); and 2) to further understand the impact of irregular sleep duration on more extreme BMI changes, we created a dichotomized variable comparing the top 5% percentile of BMI change (> 4.43 kg/m^2^) versus others, and fitted the same models.

We conducted subgroup analyses, using model 1, according to dichotomized age (>65 and ≤65), sex (female, male), and dichotomized baseline BMI (cutoff: mean baseline BMI = 27 kg/m^2^). The statistical significance of effect modification was tested on a multiplicative scale by including an interaction term between the exposure (continuous version) and specified covariates. We modeled age and BMI as continuous variables when calculating *p*-interaction.

Lastly, as an exploratory step, we analyzed the PRS-sleep interaction on the incident obesity, adjusting for covariates in model 1 and the first ten ancestry principal components to account for population stratification. This interaction was evaluated on the multiplicative scale by including an interaction term between continuous sleep duration SD and BMI PRS in model 1. Given that the BMI PRS was developed and assessed only using European ancestry White population, this analysis was conducted only among participants who identified as White, with non-missing BMI PRS data, and a baseline BMI of <30 kg/m^2^ (N = 7,551).

We used Python for data processing and visualization, and SAS version 9.4 (Cary, NC, USA) for statistical analyses. UKB Research Analysis Platform was used for aggregating UKB data including primary care entity. Our reporting aligns with guidelines provided in the Strengthening the Reporting of Observational Studies in Epidemiology (STROBE) (47).

## RESULTS

At the accelerometer study, baseline mean (SD) age was 63 (SD: 8) years. Around 52% of participants were women, 97% were White, 41% had a college degree, and 8% were employed with regular shift work occupations. Approximately 14% of participants recorded a sleep duration SD ≤30 minutes (i.e., the category for the most regular sleep duration) and 30% had a sleep duration SD exceeding 60 minutes (i.e., the category for the most irregular sleep duration; Supplemental Figure 2). Participants with higher sleep duration SD tended to be younger, more frequently women, and from non-White backgrounds, with a higher Townsend deprivation index suggesting lower socioeconomic status. Irregular sleepers were more likely to have shift work employment, current smoking, and lower physical activity levels as measured through accelerometers. Moreover, these individuals were more likely to have depression, diabetes, sleep apnea, insomnia, an evening chronotype, and daytime sleepiness, and more frequently presented with shorter accelerometer-measured sleep duration, prevalent obesity, and a higher baseline BMI (Table 1).

**Table 1.** Participants characteristics at baseline, N = 10,572.

|  |  | Sleep duration standard deviation (minutes) |  |  |  |
| --- | --- | --- | --- | --- | --- |
|  | Total<br>N = 10,572 | ≤30<br>N = 1,523 | 31-45<br>N = 3,138 | 46-60<br>N = 2,744 | >60<br>N = 3,167 |
| Age (years), mean (SD) | 63.4 (7.6) | 64.6 (7.4) | 64.1 (7.5) | 63.4 (7.5) | 62 (7.8) |
| Male % | 47.5 | 51.2 | 49.0 | 46.9 | 44.9 |
| Townsend deprivation index, mean (SD) * | -1.7 (2.8) | -2 (2.7) | -1.9 (2.6) | -1.7 (2.9) | -1.4 (2.9) |
| College or University degree % * | 40.5 | 41.3 | 41.9 | 40.6 | 38.7 |
| White % | 97.1 | 98.0 | 97.8 | 96.6 | 96.4 |
| Current Occupation/shift work % * |  |  |  |  |  |
| Employed, never/rarely shift work | 43.5 | 39.1 | 41.0 | 46.0 | 45.9 |
| Employed, sometimes/usually/always shift work | 7.7 | 5.4 | 6.6 | 6.8 | 10.6 |
| Retired/Other | 48.8 | 55.4 | 52.4 | 47.2 | 43.5 |
| Smoking status % * |  |  |  |  |  |
| Never | 55.1 | 55.4 | 55.4 | 55.2 | 54.4 |
| Previous | 38.3 | 38.3 | 38.9 | 38.6 | 37.5 |
| Current | 6.6 | 6.2 | 5.8 | 6.2 | 8.1 |
| Moderate-vigorous physical activity (min/day), mean (SD) | 38.5 (34.1) | 41.5 (34.9) | 40.5 (35.7) | 37 (32.9) | 36.2 (32.7) |
| Healthy diet score, mean (SD) *† | 3.1 (1.2) | 3.2 (1.2) | 3.1 (1.2) | 3.1 (1.2) | 3 (1.3) |
| Alcohol consumption % * |  |  |  |  |  |
| Never | 3.3 | 3.0 | 3.5 | 3.5 | 3.1 |
| Previous | 3.4 | 3.0 | 3.1 | 3.1 | 4.2 |
| Current | 93.3 | 94.0 | 93.4 | 93.3 | 92.7 |
| Dyslipidemia % | 32.4 | 32.8 | 33.5 | 32.3 | 31.3 |
| Hypertension % | 42.2 | 43.7 | 42.1 | 41.3 | 42.5 |
| Self-reported depression % * | 36.6 | 31.5 | 33.9 | 38.2 | 40.6 |
| Type 2 diabetes | 11.4 | 9.6 | 10.8 | 11.6 | 12.8 |
| Sleep apnea % | 1.7 | 0.8 | 1.4 | 1.7 | 2.2 |
| Insomnia % * |  |  |  |  |  |
| Never/rarely | 24.1 | 25.6 | 24.3 | 22.8 | 24.4 |
| Sometimes | 46.6 | 48.7 | 47.3 | 47.5 | 44.2 |
| Usually | 29.2 | 25.7 | 28.4 | 29.7 | 31.4 |
| Objective sleep duration, mean (SD) | 7.3 (0.9) | 7.5 (0.8) | 7.4 (0.8) | 7.3 (0.8) | 7 (1) |
| Chronotype % * |  |  |  |  |  |
| Definitely a 'morning' person | 23.8 | 21.5 | 24.4 | 24.4 | 23.7 |
| More a 'morning' than an 'evening' person | 33.4 | 36.8 | 34.2 | 33.5 | 30.8 |
| Intermediate | 9.6 | 10.4 | 9.8 | 9.4 | 9.3 |
| More an 'evening' than a 'morning' person | 25.1 | 24.7 | 24.8 | 25.0 | 25.6 |
| Definitely an 'evening' person | 8.1 | 6.6 | 6.8 | 7.8 | 10.5 |
| Daytime sleepiness / sleeping % * |  |  |  |  |  |
| Never/rarely | 76.3 | 77.7 | 76.7 | 76.3 | 75.4 |
| Sometimes | 20.7 | 20.6 | 20.6 | 20.5 | 21.1 |
| All of the time/Often | 3.0 | 1.7 | 2.7 | 3.2 | 3.5 |
| Baseline obesity (BMI ≥30) % | 24.9 | 19.0 | 22.3 | 26.0 | 29.4 |
| Baseline BMI (kg/m<sup>2</sup>), mean (SD) | 27.3 (5.1) | 26.5 (4.7) | 26.9 (5) | 27.5 (5.1) | 27.9 (5.4) |
| Follow-up BMI (kg/m<sup>2</sup>), mean (SD) | 27.3 (5.2) | 26.5 (4.7) | 26.9 (5) | 27.5 (5.2) | 28.1 (5.4) |
| Days between BMI measurements, mean (SD) | 896.2 (615.7) | 904.5 (619.1) | 878.6 (603.3) | 887 (604.1) | 917.6 (635.5) |

\* For variables with an asterisk, on average, the covariate assessment occurred 5 years before the conclusion of the accelerometer study.
† Healthy diet score was adopted from the American Heart Association guidelines and was created following Rutten-Jacobs et al., 2018 paper (39, 63). The table was constructed after missing values had been filled in; No single covariate had more than 2.7% missing values.
BMI: body mass index

The mean (SD) BMI at baseline was 27.3 (5.1) kg/m^2^ and 27.3 (5.2) kg/m^2^ at follow-up. These two measurements were on average 896 days apart (Q1: 441, median: 732, Q3: 1,105 days). Baseline BMI was on average measured 14 days after the accelerometer study date (Supplemental Table 4).

### Sleep duration SD and standardized BMI change

The mean for the BMI change, standardized to three-year intervals, was 0.03 kg/m^2^ (median: 0.06, Q1: -1.04, Q3: 1.27 kg/m^2^). Compared to participants with a sleep duration SD of ≤30 minutes, the standardized BMI change (95% CI) was 0.06 (-0.11, 0.22) kg/m^2^ higher for participants with sleep duration SD 31-45 minutes, 0.22 (0.05, 0.38) kg/m^2^ higher for 46-60 minutes, and 0.24 (0.08, 0.40) kg/m^2^ higher for those with >60 minutes of sleep duration SD, after adjusting for age, sex, ethnic background, Townsend deprivation index, education, occupation/shift work, and baseline BMI in model 1 (*p*-trend = 0.02; Table 2). The BMI change increased 0.12 (95%: 0.02, 0.21) kg/m^2^ for every hour longer sleep duration SD. These associations were attenuated but remained overall consistent after further adjusting for lifestyle and comorbidities in model 2 (*p*-trend = 0.02), and other sleep-related variables including sleep duration in model 3 (*p*-trend = 0.07), some of which could have a role in mediating the relationship between sleep irregularity and incident obesity.

**Table 2.** Longitudinal association between irregular sleep duration and BMI change measures, N = 10,572.

|  |  |  |  |  |  |
| --- | --- | --- | --- | --- | --- |
| Exposure | Mean (SD) for outcome variable | Sample size | Beta (95% CI) Model 1<sup>†</sup> | Beta (95% CI) Model 2<sup>‡</sup> | Beta (95% CI) Model 3<sup>§</sup> |
| Sleep duration SD and BMI change (kg/m<sup>2</sup>) |  |  |  |  |  |
| ≤30 mins | -0.07 (2.56) | 1523 | Ref. | Ref. | Ref. |
| 31-45 mins | -0.04 (2.62) | 3138 | 0.06 (-0.11, 0.22) | 0.06 (-0.10, 0.22) | 0.05 (-0.11, 0.21) |
| 46-60 mins | 0.09 (2.72) | 2744 | **0.22 (0.05, 0.38)** | **0.21 (0.05, 0.38)** | **0.20 (0.03, 0.37)** |
| >60 mins | 0.11 (2.78) | 3167 | **0.24 (0.08, 0.40)** | **0.23 (0.06, 0.39)** | **0.20 (0.03, 0.37)** |
| Per 1-hour increment | 0.03 (2.69) | 10572 | **0.12 (0.02, 0.21)** | **0.11 (0.02, 0.21)** | 0.09 (-0.01, 0.19) |
| *p*-trend* | NA | NA | **0.0155** | **0.0212** | 0.0717 |
| Sleep duration SD and relative BMI change percentage |  |  |  |  |  |
| ≤30 mins | -0.05 (9.49) | 1523 | Ref. | Ref. | Ref. |
| 31-45 mins | 0.09 (9.41) | 3138 | 0.22 (-0.36, 0.80) | 0.23 (-0.35, 0.80) | 0.20 (-0.38, 0.77) |
| 46-60 mins | 0.55 (9.71) | 2744 | **0.79 (0.19, 1.38)** | **0.78 (0.18, 1.37)** | **0.73 (0.13, 1.32)** |
| >60 mins | 0.58 (9.62) | 3167 | **0.81 (0.23, 1.40)** | **0.77 (0.19, 1.36)** | **0.68 (0.09, 1.27)** |
| Per 1-hour increment | 0.34 (9.56) | 10572 | **0.39 (0.05, 0.73)** | **0.37 (0.03, 0.71)** | 0.31 (-0.05, 0.66) |
| *p*-trend* | NA | NA | **0.0231** | **0.0309** | 0.0878 |
| * Continuous sleep duration SD was included when calculating *p*-trend.<br>† Model 1 adjusted for age in years, sex, ethnic background, Townsend deprivation index, education, occupation/shift work, and baseline BMI<br>‡ Model 2 adjusted for covariates in model 1 along with smoking, physical activity, healthy diet score, alcohol consumption, dyslipidemia, hypertension, depression, and diabetes.<br>§ Model 3 adjusted for covariates in model 2 along with sleep apnea, insomnia, accelerometer measured sleep duration, chronotype, and daytime sleepiness.<br>Both BMI change outcomes were standardized to three-year intervals.<br>Results from linear regression models.<br>SD: Standard deviation, BMI: Body mass index, CI: confidence interval, NA: not applicable |  |  |  |  |  |

In the sensitivity analysis evaluating extreme BMI change based on the dichotomized outcome of at least 4.43 kg/m^2^ BMI change, across all models we observed a similar pattern with participants with 46-60 minutes of sleep duration SD having increased risk for extreme BMI gain compared to those with ≤30 minutes [comparative RR (95% CI) in model 1: 1.41 (1.04, 1.91), *p*-trend: 0.2180, Supplemental Table 5]. When comparing sleep duration SD of >60 to ≤30 minutes, this association was only significant in model 1 [RR (95% CI): 1.35 (1.003, 1.83)].

### Sleep duration SD and relative BMI change percentage

The mean for relative BMI change percentage was 0.34% (Median: 0.23%, Q1: -3.91%, Q3: 4.81%). As baseline sleep duration SD increased, the mean for BMI change percentage during follow-up increased. In model 1, compared to participants with a sleep duration SD of ≤30 minutes, the relative BMI change (95% CI) was 0.22% (-0.36, 0.80) higher for those with a sleep duration SD 31-45 minutes, 0.79% (0.19, 1.38) higher for 46-60 minutes, and 0.81% (0.23, 1.40) higher for >60 minutes of sleep duration SD (*p*-trend = 0.02; Table 2). For every hour increase in sleep duration SD, the relative BMI change increased by 0.39% (95% CI: 0.05, 0.73). The associations were slightly attenuated and remained statistically significant after adjustment for lifestyle factors, along with metabolic comorbidities and depression in model 2 (*p*-trend = 0.03), and after further adjusting for sleep-related covariates in model 3 (*p*-trend = 0.09).

### Sleep duration SD and incident obesity

Among participants with a baseline BMI of <30 kg/m^2^ (N = 7,937), a total of 383 developed obesity (BMI ≥30 kg/m^2^). The follow-up period between the two measurements was on average 2.5 years (median = 2; interquartile range = 1.8 years). In model 1, compared to participants with a sleep duration SD of ≤30 minutes, the covariate-adjusted RR (95% CI) for incident obesity was 1.12 (0.79, 1.60) for 31-45 minutes, 1.49 (1.05, 2.11) for 46-60 minutes, and 1.80 (1.28, 2.52) for >60 minutes of sleep duration SD (*p*-trend = 0.0028; Table 3). In model 2, which included additional covariates that may be on the mediating pathway between sleep and BMI, or downstream of these two (i.e., collider), the associations weakened and remained statistically significant when comparing participants with the highest level of sleep irregularity (sleep duration SD >60 minutes) to those with a sleep duration SD of ≤30 minutes (*p*-trend = 0.01). In model 3, the strength of this association showed a slight decrease, but continued to be statistically significant [RR (95% CIs) comparing sleep duration SD >60 to ≤30 mins: 1.60 (1.14, 2.24); *p*-trend = 0.09]. The E value (lower 95% limit) for this association was 2.58 (1.54). This suggests that the observed association could be explained by an unknown or unmeasured confounder associated with both sleep duration SD and incident obesity by an RR as large as 2.58 (1.54), conditional on covariates in model 3. This moderate E value implies that the observed association is unlikely to be explained by the existence of a strong unobserved confounder, particularly considering the wide range of covariates included in model 3.

**Table 3.** Associations between irregular sleep duration and risk for incident obesity in participants without obesity at baseline, N = 7,937.

| Exposure | Incident Obesity<br>/ Sample Size | RR (95% CI)<br>Model 1<sup>†</sup> | RR (95% CI)<br>Model 2<sup>‡</sup> | RR (95% CI)<br>Model 3<sup>§</sup> |
| --- | --- | --- | --- | --- |
| Sleep duration SD categories |  |  |  |  |
| ≤30 mins | 42 / 1233 | Ref. | Ref. | Ref. |
| 31-45 mins | 93 / 2437 | 1.12 (0.79, 1.60) | 1.11 (0.78, 1.58) | 1.10 (0.77, 1.57) |
| 46-60 mins | 104 / 2030 | **1.49 (1.05, 2.11)** | **1.43 (1.01, 2.02)** | 1.40 (0.99, 1.98) |
| >60 mins | 144 / 2237 | **1.80 (1.28, 2.52)** | **1.69 (1.21, 2.36)** | **1.60 (1.14, 2.24)** |
| Per 1-hour increment | 383 / 7937 | **1.24 (1.08, 1.42)** | **1.21 (1.05, 1.39)** | 1.15 (0.98, 1.34) |
| *p*-trend* | NA | **0.0028** | **0.0100** | 0.0864 |

\* Continuous irregular sleep duration standard deviation was included when calculating *p*-trend.
† Model 1 adjusted for age in years, sex, ethnic background, Townsend deprivation index, education, occupation/shift work, and follow-up period (days between the two BMI measurements)
‡ Model 2 adjusted for covariates in model 1 along with smoking, physical activity, healthy diet score, alcohol consumption, dyslipidemia, hypertension, depression, and diabetes.
§ Model 3 adjusted for covariates in model 2 along with sleep apnea, insomnia, accelerometer measured sleep duration, chronotype, and daytime sleepiness.
Results from Poisson regression models with a robust error variance. Participants with a baseline BMI $\geq 30$ kg/m<sup>2</sup> were excluded from analyses reported in this table.
SD: Standard deviation, BMI: Body mass index, RR: risk ratio, CI: confidence interval, NA: Not applicable

### Sub-group analysis (effect heterogeneity)

We did not observe evidence for differences in associations in subgroup analyses by age, sex, or baseline BMI (*p-*interaction ≥0.29; Table 4), although stronger and statistically significant positive associations were more consistently observed among participants aged 65 years or younger across all BMI change outcomes.

**Table 4.** Associations between irregular sleep duration (per 1-hour increment in SD) and BMI change outcomes stratified by age, sex, and baseline BMI.

|  |  |  |  |  |  |  |
| --- | --- | --- | --- | --- | --- | --- |
|  | BMI change (kg/m<sup>2</sup>)<br>N = 10,572 * |  | Relative BMI change percentage<br>N = 10,572 * |  | Incident obesity<br>N = 7,937 † |  |
|  | Beta (95% CI) | *p*-interaction | Beta (95% CI) | *p*-interaction | RR (95% CI) | *p*-interaction |
| Age |  |  |  |  |  |  |
| Age >65 | 0.06 (-0.09, 0.20) | 0.9728 | 0.19 (-0.34, 0.71) | 0.8498 | 1.14 (0.92, 1.40) | 0.8310 |
| Age ≤65 | **0.16 (0.04, 0.29)** |  | **0.54 (0.10, 0.99)** |  | **1.31 (1.09, 1.57)** |  |
| Sex |  |  |  |  |  |  |
| Female | 0.09 (-0.05, 0.22) | 0.5056 | 0.31 (-0.18, 0.79) | 0.5840 | **1.24 (1.02, 1.50)** | 0.9666 |
| Male | **0.14 (0.01, 0.28)** |  | 0.47 (-0.003, 0.94) |  | **1.23 (1.01, 1.51)** |  |
| Baseline BMI |  |  |  |  |  |  |
| Baseline BMI >27 | 0.14 (-0.03, 0.30) | 0.2892 | 0.44 (-0.07, 0.95) | 0.2878 | 1.12 (0.96, 1.30) | 0.8687 |
| Baseline BMI ≤27 | 0.10 (-0.004, 0.21) |  | 0.38 (-0.07, 0.83) |  | 1.15 (0.71, 1.88) |  |
| * The continuous outcomes were standardized to three-year intervals. Models were adjusted for age in years, sex, ethnic background, Townsend deprivation index, education, occupation/shift work, and baseline BMI.<br>† Adjusted for age in years, sex, ethnic background, Townsend deprivation index, education, occupation/shift work, and follow-up period (days between the two BMI measurements). Participants with a baseline BMI >30 were excluded from analyses for incident obesity outcome.<br>Interaction was tested on the multiplicative scale.<br>Continuous variables, including age (in years), sleep duration SD, and baseline BMI, were used in calculating *p*-interactions and adjusting for residual confounding in subgroup analyses.<br>SD: Standard deviation, BMI: Body mass index, RR: risk ratio, CI: confidence interval |  |  |  |  |  |  |

We did not identify a BMI PRS-sleep SD interaction on risk for incident obesity (*p-* interaction in model 1: 0.88; Table 5). However, in stratified analyses, the associations between sleep duration SD and risk for incident obesity was statistically significant only among participants with lower genetic predisposition to higher BMI as measured by the BMI PRS. The RR (95% CI) for incident obesity associated with 1-hour increase in sleep duration SD was 1.43 (1.13, 1.81) in participants with low PRS, 1.15 (0.90, 1.46) in intermediate PRS, and 1.20 (0.95, 1.51) in high PRS group.

**Table 5.** Associations between irregular sleep duration (per 1-hour increment in SD) and Incident obesity stratified by BMI polygenic risk score, N = 7,551.

|  | Incident obesity cases / Sample size | RR (95% CI) | *P*-interaction |
| --- | --- | --- | --- |
|  |  |  | 0.8758 |
| Low BMI PRS ($\leq 33^{\text{rd}}$ percentile; PRS $\leq -0.617$) | 105 / 2864 | **1.43 (1.13, 1.81)** | |
| Intermediate BMI PRS ($33^{\text{rd}} - 66^{\text{th}}$ percentile; PRS -0.618 to 0.207) | 118 / 2519 | 1.15 (0.90, 1.46) | |
| High BMI PRS ($> 66^{\text{th}}$ percentile; PRS $> 0.207$) | 138 / 2168 | 1.20 (0.95, 1.51) | |
| Adjusted for age in years, sex, Townsend deprivation index, education, occupation/shift work, follow-up period (days between the two BMI measurements), and first 10 genetic principal components.<br>Participants with a baseline BMI $\geq 30$ kg/m<sup>2</sup>, non-white, or missing BMI PRS information were excluded from the analyses in this table. Interaction on the multiplicative scale was tested using continuous sleep duration SD and continuous PRS.<br>BMI: Body mass index, PRS: Polygenic risk score, RR: risk ratio, CI: confidence interval | | | |

## DISCUSSION

In a large longitudinal study of middle-aged to older adults in the UK, we found that irregular sleep duration, quantified by 7-day sleep duration SD, was associated with greater BMI change and an increased risk for incident obesity over an average follow-up of 2.5 years. These associations persisted even after adjusting for a comprehensive set of confounders, including sociodemographic and lifestyle factors, comorbidities, and other sleep-related covariates, such as average sleep duration, some of which might be on mediating pathways (e.g., dyslipidemia). Age, sex, baseline BMI, and genetic predisposition to higher BMI did not modify these associations; however, the associations were more pronounced among younger participants and those with a lower genetic risk of BMI.

The observed associations between irregular sleep duration SD and BMI change outcomes were modest in magnitude. However, these associations are comparable to other observational studies evaluating lifestyle and weight change (48-50). For example, in a large cohort study of 120,877 U.S. men and women, the weight change over a 4-year period was 3.93 pounds (1.78 kg) more comparing extreme quintiles of dietary changes (i.e., the worst versus the best dietary score changes) and 1.76 pounds (0.80 kg) more comparing extreme quintiles of physical activity changes (i.e., the lowest versus the highest changes in physical activity levels) (48). For an individual of 175 cm height, these estimates corresponded to a difference of 0.58 kg/m^2^ and 0.26 kg/m^2^, respectively, in BMI changes, comparable to our observation of 0.24 kg/m^2^ more BMI change over an average of 2.5 years comparing individuals with sleep duration SD >60 min versus ≤30 min. Longer follow-up for long-term weight change might allow us to observe stronger associations, as the effect of irregular sleep patterns on metabolic health may accumulate over time. Nonetheless, even small reductions in BMI through improvements in sleep regularity could have important public health implications, especially given the widespread prevalence of sleep irregularity as observed in our study (e.g., 30% with sleep duration SD >60 min) and the strength of this association, which is comparable to other established lifestyle factors like diet and physical activity. Moreover, in high-risk groups, such as those with pre-diabetes, lifestyle interventions often outperform medications in prevention of relevant metabolic diseases and remain a crucial component in treatment plans (51).

Three published systematic reviews have evaluated some measure of sleep variability, assessed via different tools and indices, and body composition outcomes, together covering more than 50 papers (9, 17, 52). Most studies employed a cross-sectional design, which limits the ability to discern the temporal relationship between irregular sleep and changes in adiposity. The majority of cross-sectional studies identified a significant positive association, with fewer reporting null results (52). The largest cross-sectional study to-date, involving 120,522 adults, found an association between sleep duration variability, as measured by the sleep duration SD captured using Fitbit wearable devices over at least 100 nights, and increased self-reported BMI (19).

Only a limited number of longitudinal studies have evaluated the association between irregular sleep duration and BMI change outcomes, with even fewer focusing on middle-aged to older adults (9, 17, 52). A recent panel review confirmed the importance of sleep regularity for overall and metabolic health, noting the need for more high-quality, prospective studies (9). Similar to our findings, a recent relatively large prospective analysis, involving 6,785 participants from the All of Us Research Program with a median sleep monitoring period of 4.5 years, identified a positive association between higher accelerometer-measured sleep duration SD and incident obesity [Hazard Ratio (95% CI) comparing 75^th^ to the 25^th^ percentile of sleep duration SD: 1.21 (1.08–1.37)] (24). Another smaller prospective analysis, involving 1,986 overweight and obese adults in Spain with metabolic syndrome enrolled in a 12-month weight loss trial, identified a positive association between higher accelerometer-measured sleep duration SD and increased BMI, specifically in relation to a poorer response to the weight loss intervention (25).

Interestingly, we found stronger and more consistent associations between irregular sleep duration and BMI change among younger participants. Middle age is the life period most susceptible to weight gain. In contrast, weight loss is more common in older individuals due to muscle mass loss, which may limit our ability to examine the association with weight gain in this group. Future studies may further investigate the association across different age groups and explore the development and assessment of interventions to improve sleep regularity as a strategy to reduce obesity. Additionally, we did not observe a BMI PRS-sleep SD interaction, although the association between sleep duration irregularity and risk for incident obesity was statistically significant only among individuals with lower genetic predisposition to obesity. However, this analysis was exploratory, and both the sample size and the number of cases were smaller than in the primary analyses. Nevertheless, the point estimates for irregular sleep duration and the incident obesity associations were consistently above 1 across low, intermediate, and high BMI PRS groups.

There are two widely recognized theories on the development of obesity: the energy balance model, which posits that a positive energy balance (where calorie intake exceeds expenditure) leads to net fat deposition; and the carbohydrate-insulin model, which suggests that altered fuel partitioning (where energy is directed toward storage rather than use) shifts energy storage towards fat deposition, ultimately contributing to obesity (53). The mechanism through which irregular sleep patterns increases risk of obesity could be explained under both theories (53). Irregular sleep duration may adversely influence sleep quantity and sleep quality and is correlated with metabolically unhealthy sleep-related traits such as evening chronotype, which collectively lead to energy imbalance and weight gain (54-57). Further, irregular sleep duration can disrupt the circadian timing system that regulates metabolic functions by interfering with external cues, such as light, which in turn affects glucose metabolism, insulin secretion, and appetite-regulating hormones, as well as causing misalignment between biological pathways driven by multiple peripheral and central clocks and external behaviors, adversely impacting metabolism (6, 54). Moreover, irregular sleep duration may contribute to other metabolically unhealthy behaviors, such as late-night snacking, breakfast skipping, poor dietary choices, and sedentary behaviors, leading to weight gain and increased risk of obesity (58). In our analyses, adjusting for lifestyle and sleep-related factors only modestly attenuated the association between sleep duration SD and BMI change outcomes. Further research, using other measures of fat mass, may help elucidate the mechanisms underlying the observed association (9).

Study strengths include a longitudinal design with a large sample, use of validated device-based sleep data collected in habitual settings, objective measurement of BMI and physical activity, and adjustments for a wide range of comorbidities and lifestyle factors. We also utilized a sleep variability metric that simplifies interpretations compared to other measures like the sleep regularity index, allowing for actionable public health recommendations to improve sleep regularity (9, 59).

While use of objectively measured BMI had many advantages over self-reported BMI as in many prior studies, there are some concerns regarding the BMI change outcome in our study. First, we estimated BMI change using two BMI measurements, assuming a linear change, which may not fully capture the complexity and variability of weight changes over time. This simplified measure of BMI change could introduce misclassification, potentially attenuating the observed associations. Second, weight and BMI data from primary care records may be prone to measurement errors since they were collected in non-research settings. Third, BMI is a metric for general obesity and does not reflect the distributions of body fat or lean mass. Specifically, BMI may underestimate body fat in older individuals due to age-related muscle loss potentially introducing bias in our results (60).

Our study has some other limitations. Notably, intervals between assessments of certain confounders and sleep duration SD measurements spanned 0 to 9 years (median: 5 years). These gaps may not impact stable variables like demographics but could introduce biases in lifestyle confounders that change over time such as smoking status, diet, and alcohol use. To reduce potential residual confounding, we updated confounder data for participants attending follow-up visits post-baseline (n = 1,455), using the assessment closest to the accelerometer study date. E-values also suggested minimal risk for unmeasured confounding. Another limitation concerns the measurement of sleep duration SD from a 7-day period, which might not reflect the long-term habitual sleep patterns of some participants. The response rate for the UKB was approximately 5.5% (27) and the selection of participants for the accelerometer study was conducted through a constrained random sampling method, which may increase the risk of selection bias (61). Despite this, the associations between risk factors and outcomes observed in the UKB data closely matched those from other representative studies with standard response rates, suggesting our findings may be generalizable to larger populations (62). Nonetheless, participants in our study were primarily healthy individuals who identified as White, limiting the generalizability to other groups. Lastly, the lack of data on intentional weight loss, such as those interventions through anti-obesity medications or lifestyle changes, limits our understanding of their potential role as effect modifiers in the observed associations.

In summary, in a large prospective study with a mean follow-up of 2.5 years among middle-aged to older adults, we found that greater sleep duration SD, indicative of irregular sleep patterns, was associated with a modest increase in standardized and relative BMI change and a higher risk of incident obesity compared to those with more regular sleep durations. These associations largely persisted, with similar magnitude, even after adjusting for sociodemographic and lifestyle factors, comorbidities, and sleep-related covariates. Given the high prevalence of irregular sleep duration, these findings highlight the potential for clinical trials testing interventions that target sleep irregularity to reduce obesity. With favorable results, sleep regularity could become part of public health strategies tackling obesity.

## Data availability

This study used data from the UK Biobank under approved Project ID: 85501. UK Biobank data are available upon application to registered researchers (https://www.ukbiobank.ac.uk).

## Code availability

The code used to generate the results is available upon request from the first author (SK).

## Contributions

SK and TH contributed to the conception, investigation, data curation, methodology, and formal analysis of the project. SK prepared the original draft of the manuscript, and all authors reviewed, revised, participated in data interpretation, and approved the final version of the manuscript. TH and SR supervised the work.

## Acknowledgments

The authors express their gratitude to the participants and staff of the UK Biobank study for their invaluable contributions of data and support to this research. The authors also acknowledge the support of all funding sources that facilitated this study.

