## Supplemental for "Association between Accelerometer-Measured Irregular Sleep Duration and Longitudinal Changes in Body Mass Index in Older Adults"

Supplemental Text: Primary care data cleaning and BMI change equations

In the primary care dataset, Read v2 and CTV3 codes, specifically '22A..' for weight measured during examination and '22K..' for BMI, were utilized to retrieve relevant measurements. Primary care data have been minimally cleaned. Following the distribution of weight and BMI measurements from the UK Biobank baseline visit, we excluded primary care weight values of less than 30 or more than 198 kg. Similarly, we excluded BMI measurements less than 12 or more than 75 kg/m$^2$. These cutoffs were selected based on the minimum and maximum values of corresponding measurements from baseline center visits in the UK Biobank cohort. Further, for participants with multiple primary care weight or BMI measurements, we conducted a manual review of participant data and identified and excluded certain cases with implausible entries (e.g., sudden jump/fall in the weight values, two measurements in the same day with largely different values). We retained up to two weight or two BMI measurements from the primary care data and assessment center datasets. The first measurement (baseline) retained was the one closest to accelerometer study date, subject to the condition that it occurred no earlier than one year before and no later than one year after the accelerometer study date. The follow-up measurement chosen was the one closest to three years after the baseline measurement, provided it occurred after the baseline measurement and at least six months after the accelerometer study date. Missing BMI values were calculated based on height and weight when available. Below are the equations used for defining BMI change outcomes.

$$\boldsymbol{Standardized\ BMI\ change} = \frac{Follow\ up\ BMI - Baseline\ BMI}{\text{Time Difference in Days}} \times\ 1095\ Days$$

$$\boldsymbol{Relative\ BMI\ change} = \frac{Standardized\ BMI\ Change}{\text{Baseline BMI}} \times\ 100$$

Supplemental Figure 1. Study flow diagram

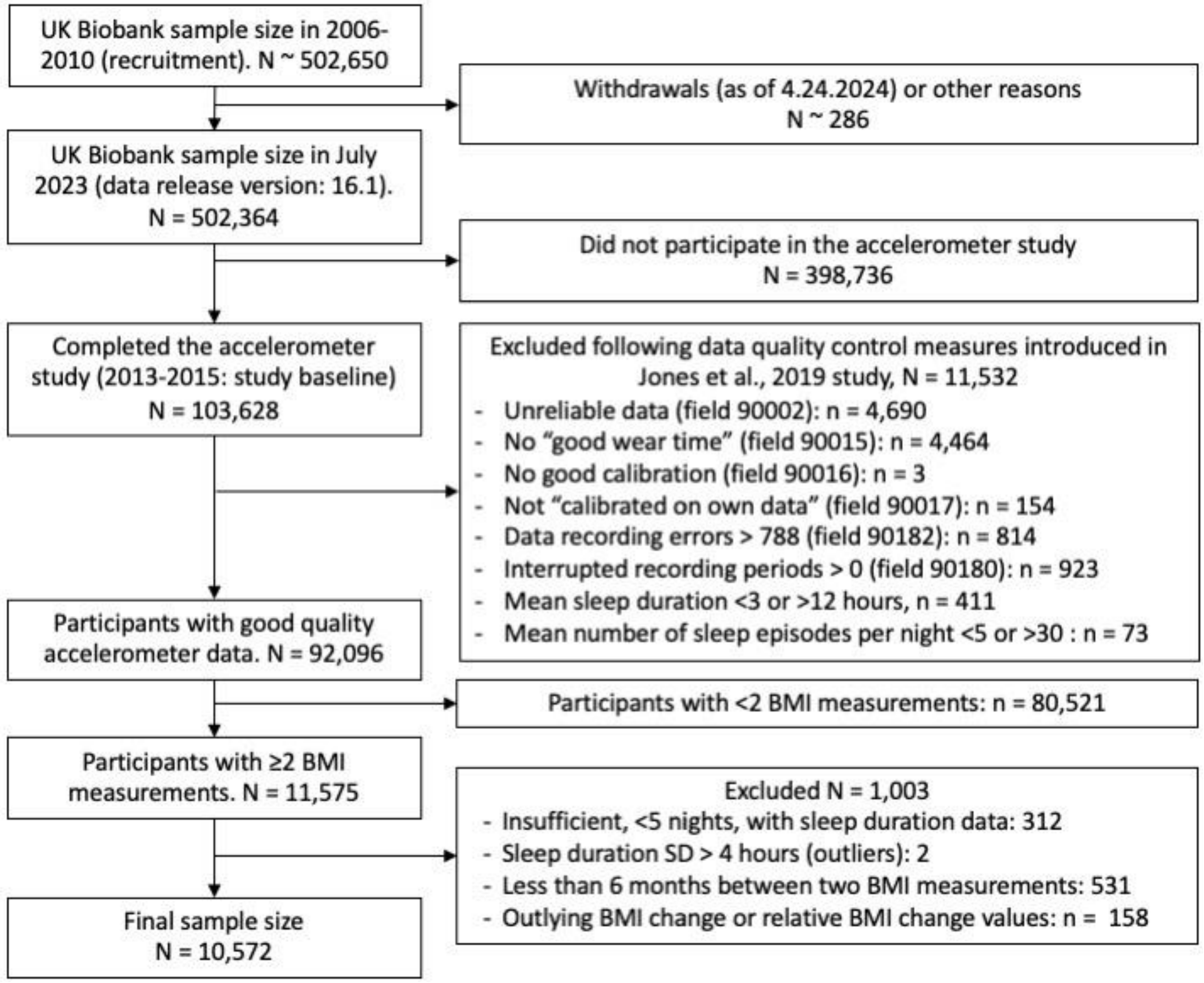

Quality control measures are available in the UK Biobank dataset.
We used sleep measures derived from accelerometer data in the Jones 2019 paper. Insufficient nights with sleep duration data, defined as having fewer than five nights recorded, were identified using a variable derived by Jones et al. (2019) named 'n_nights_sleep'.
Participants with missing values for quality control measures were also excluded from analyses.
Reference to Jones 2019 paper: Jones SE, Lane JM, Wood AR, van Hees VT, Tyrrell J, Beaumont RN, et al. Genome-wide association analyses of chronotype in 697,828 individuals provides insights into circadian rhythms. Nature communications. 2019;10(1):343.
BMI: Body mass index

Supplemental Figure 2. Distribution of sleep duration standard deviation measured across 7-day accelerometer study

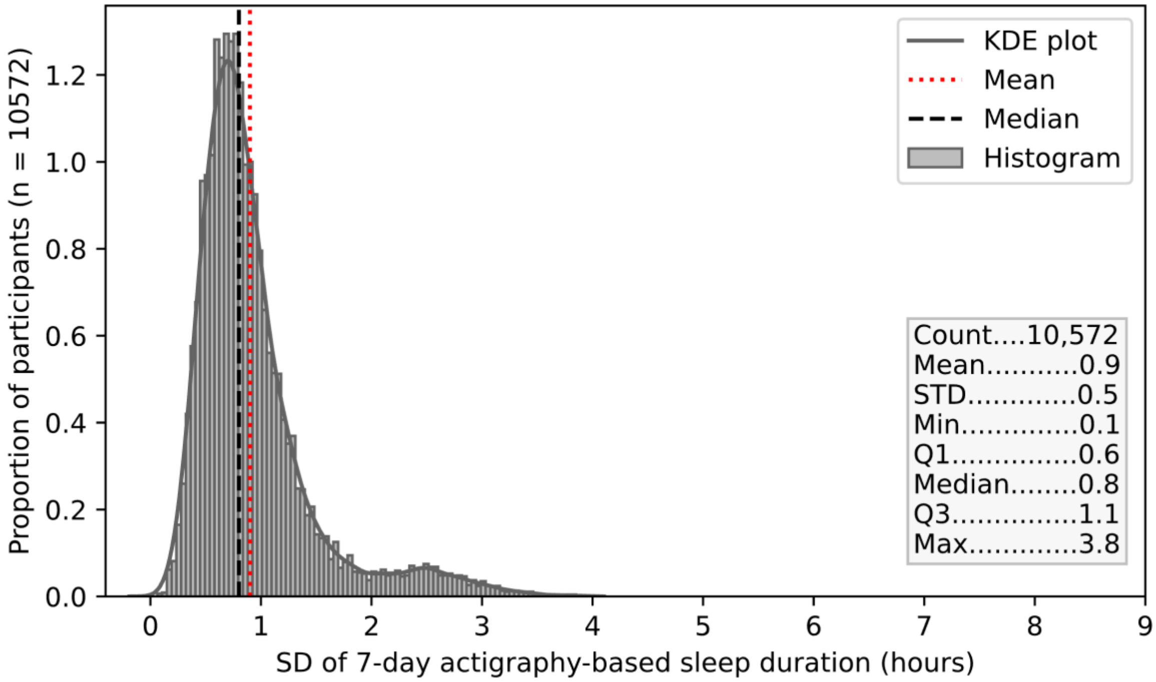

Participants with more than >4 hours of sleep duration standard deviation (outliers) were excluded from this data visualization.

Supplemental Figure 3. Distribution of BMI change outcomes (standardized to three-year intervals)

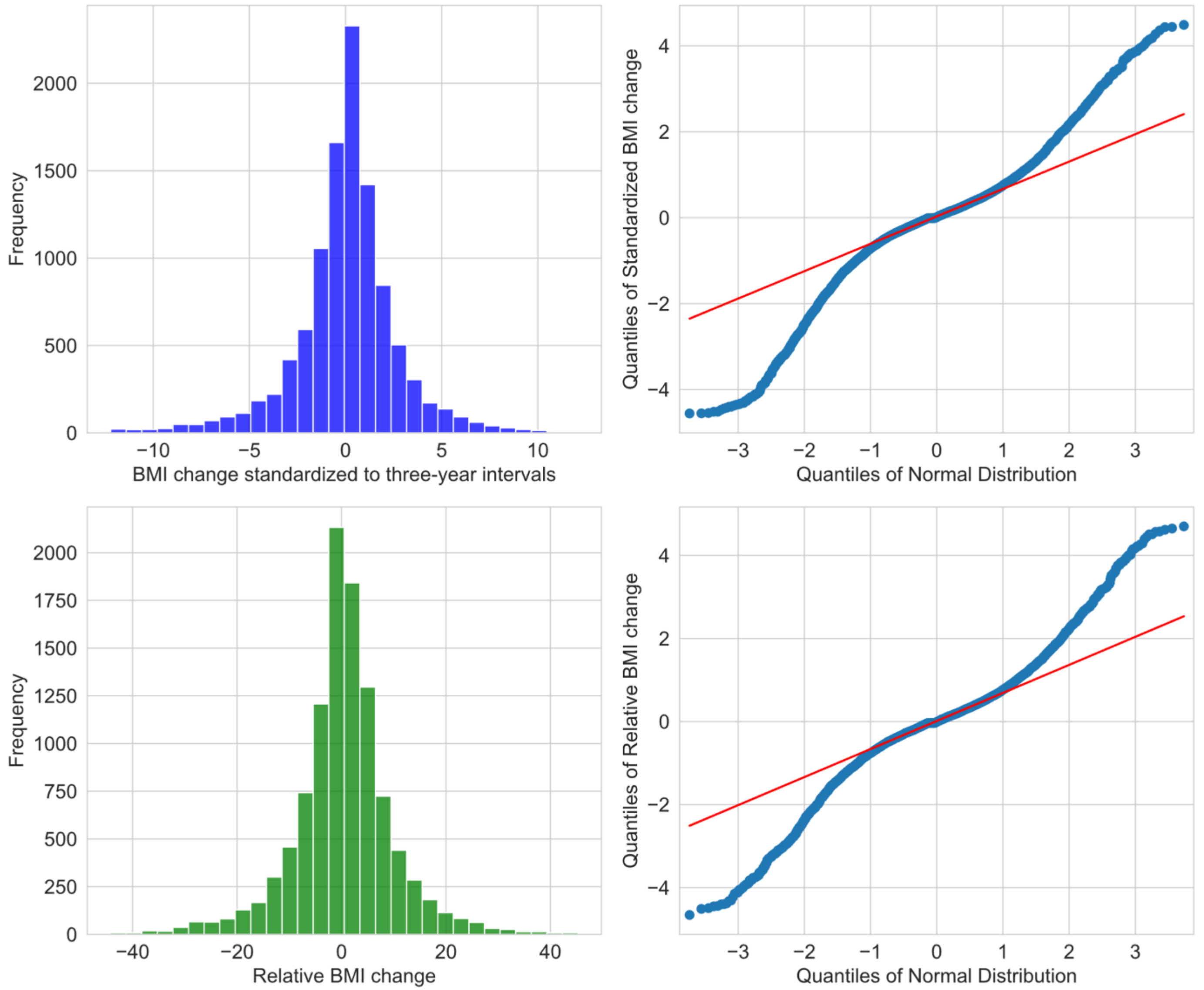

Supplemental Figure 4. Variable assessment year.

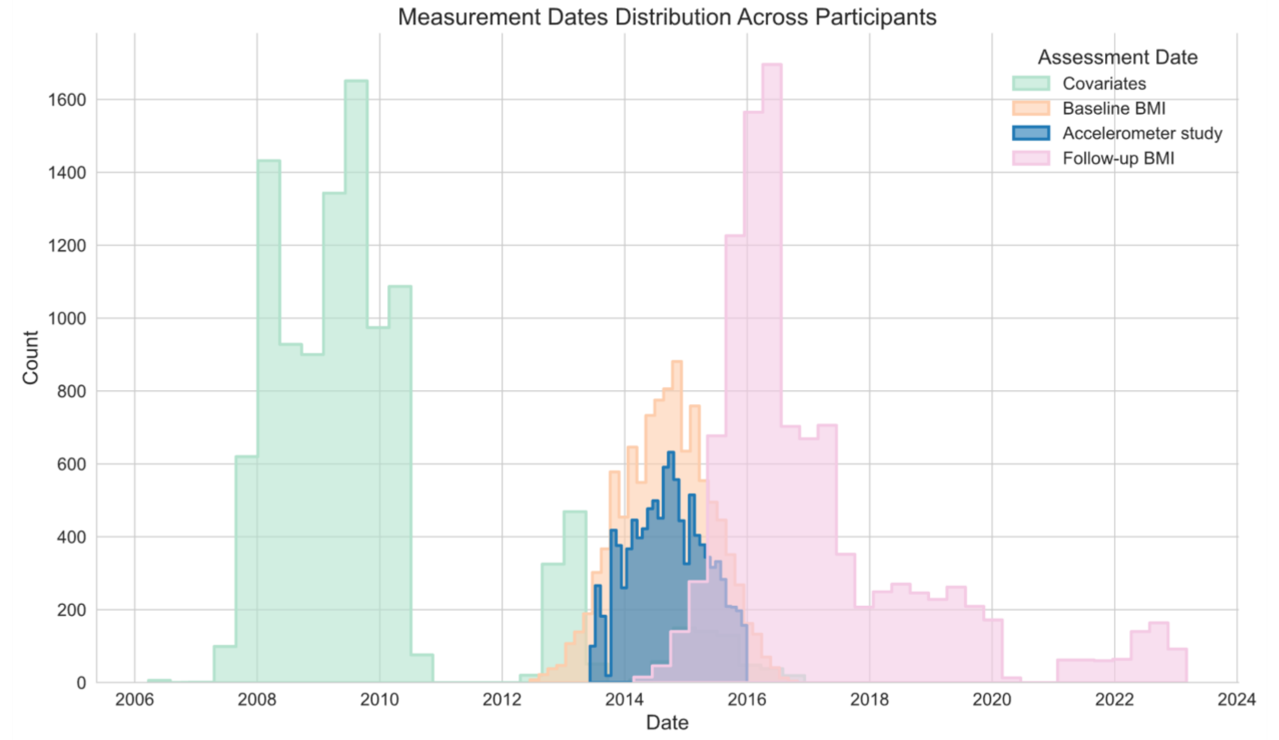

Histograms were plotted using seaborn.histplot with bins=30
For covariates (green plot), when available, we updated the measurements using data closer to the accelerometer study date, ensuring that these measurements occurred before or at most 365 days after the acccelerometer study date.

| Supplemental Table 1. Participants baseline characteristics comparing those included and excluded | | | |
|---|---|---|---|
| | **UK Biobank Accelerometry study N = 103,628** | **Included N = 10,572** | **Excluded N = 93,056** |
| **Age (years), mean (SD)** | 62.3 (7.9) | 63.4 (7.6) | 62.1 (7.9) |
| **Male %** | 43.8 | 47.5 | 43.4 |
| **Townsend deprivation index, mean (SD)** | -1.7 (2.8) | -1.7 (2.8) | -1.7 (2.8) |
| **College or University degree %** | 44.7 | 40.5 | 45.2 |
| **White %** | 96.8 | 97.1 | 96.7 |
| **Current Occupation/shift work %** | | | |
| Employed, never/rarely shift work | 49.9 | 43.5 | 50.7 |
| Employed, sometimes/usually/always shift work | 7.8 | 7.7 | 7.8 |
| Retired/Other | 42.3 | 48.8 | 41.6 |
| **Moderate-vigorous physical activity (min/day), mean (SD)** | 40.2 (35.9) | 38.5 (34.1) | 40.4 (36.1) |
| **Objective sleep duration, mean (SD)** | 7.2 (1.0) | 7.3 (0.9) | 7.2 (1.1) |
| **Weight recorded at assessment center visit, mean (SD)** | 76.7 (15.4) | 79.5 (16.6) | 76.4 (15.2) |
| **BMI recorded at assessment center visit, mean (SD)** | 26.7 (4.5) | 27.7 (5.0) | 26.6 (4.5) |
| The table was constructed after missing values had been filled in for “Included” column. No single covariate had more than 2.7% missing values. Missing values were not filled for “UK Biobank accelerometry study” and “Excluded” columns. | | | |

| Supplemental Table 2. UK Biobank Data-Field numbers of variables used in this study | | |
|---|---|---|
| **Variable** | **Data-Field** | **Notes on variable definition** |
| Body mass index | 21001, 42040 (Record Table 1060) | Read 2 and Read 3 codes "22A.." and "22K.." were used to obtain primary care BMI and weight measurements. |
| End time of accelerometer wear | 90011 | This date marks the baseline of the study, corresponding to when the exposure (i.e., sleep duration SD) was evaluated. |
| Date of attending assessment center | 53 | -- |
| Age | 34, 52 | -- |
| Sex | 31 | -- |
| Ethnic background | 21000 | -- |
| Townsend deprivation index | 22189 | -- |
| Education | 6138 | -- |
| Occupation/shift work | 826, 6142 | -- |
| Smoking status | 20116 | -- |
| Physical activity | 40049 | Following Bradbury et al., 2016 paper (1), values >6 hours per day were set to 6 hours. |
| Healthy diet score | 1289, 1299, 1309, 1319, 1329, 1339, 1349, 1359, 1369, 1379, 1389 | We followed instructions from Rutten-Jacobs et al., 2018 (2) (Text S1 and Table S1); Pazoki et al., 2018 (3); and Wang et al., 2018 (4). |
| Alcohol consumption | 20117 | -- |
| Lipidemia | 130814, 20003 | Dyslipidemia was defined based on ICD-10 code E78 or self-reported lipid-lowering medications, including simvastatin, pravastatin, fluvastatin, atorvastatin, rosuvastatin, ezetimibe, nicotinic acid product, or fenofibrate. Medications were selected following Tikkanen et al., 2018 paper (5). |
| Hypertension first occurrence | 131286, 131288, 131290, 131292, 131294, 132180 | Hypertension was based on ICD-10 codes I10-I13, I15, and O10. |
| Self-reported depression | 2090, 2100, 2050, 2060 | Self-reported depression was defined as a self-reported clinic visit for depression or a Patient Health Questionnaire score of 3 or above. |
| Type 2 Diabetes | 130708, 130709 | ICD-10 code E11 first reported at/before accelerometer study or diabetes medication use reported at assessment visit. |
| Sleep apnea | 131060, 41270 | Based on ICD-10 code G47. |
| Insomnia | 1200 | -- |
| Chronotype | 1180 | -- |
| Daytime dozing / sleeping | 1220 | -- |
| BMI Polygenic Risk Score | 26216 | -- |
| Genetic principal components | 22009 | -- |
| Accelerometer derived variables and their QC measures | Please see Return 1862 and Jones et al., 2019 (6). | Return 1862: https://biobank.ctsu.ox.ac.uk/crystal/dset.cgi?id=1862). QC measures are listed in Supplemental Figure 1 |

| Supplemental Table 3. Participants baseline characteristics comparing those with and without updated baseline measurements | | |
|---|---|---|
| | **Without updated measurement N = 9,117** | **With updated measurement (>1 instances) N = 1,455** |
| **Age (years), mean (SD)** | 63.4 (7.7) | 63.2 (7.6) |
| **Male %** | 47.2 | 49.6 |
| **Townsend deprivation index, mean (SD)** | -1.6 (2.8) | -2.1 (2.6) |
| **College or University degree %** | 39.8 | 44.7 |
| **White %** | 97.0 | 98.0 |
| **Current Occupation/shift work %** | | |
| Employed, never/rarely shift work | 44.8 | 35.2 |
| Employed, sometimes/usually/always shift work | 8.0 | 5.6 |
| Retired/Other | 47.2 | 59.2 |
| **Smoking status %** | | |
| Never | 54.5 | 58.2 |
| Previous | 38.3 | 38.4 |
| Current | 7.1 | 3.4 |
| **Moderate-vigorous physical activity (min/day), mean (SD)** | 38.3 (33.9) | 39.8 (35.2) |
| **Healthy diet score, mean (SD) †** | 3.1 (1.2) | 3.1 (1.2) |
| **Alcohol consumption %** | | |
| Never | 3.4 | 3.1 |
| Previous | 3.5 | 3.1 |
| Current | 93.2 | 93.8 |
| **Lipidemias %** | 31.8 | 36.4 |
| **Hypertension %** | 42.4 | 40.9 |
| **Self-reported depression %** | 36.8 | 35.6 |
| **Type 2 diabetes** | 11.6 | 10.2 |
| **Sleep apnea %** | 1.7 | 1.5 |
| **Insomnia %** | | |
| Never/rarely | 24.2 | 23.8 |
| Sometimes | 46.7 | 46.3 |
| Usually | 29.1 | 29.9 |
| **Objective sleep duration, mean (SD)** | 7.3 (0.9) | 7.2 (0.9) |
| **Chronotype %** | | |
| Definitely a 'morning' person | 23.5 | 25.4 |
| More a 'morning' than an 'evening' person | 33.4 | 33.2 |
| Intermediate | 9.6 | 9.8 |
| More an 'evening' than a 'morning' person | 25.5 | 22.5 |
| Definitely an 'evening' person | 8.0 | 9.1 |
| **Daytime dozing / sleeping %** | | |
| Never/rarely | 76.5 | 75.7 |
| Sometimes | 20.7 | 20.9 |
| All of the time/Often | 2.9 | 3.4 |
| **Baseline BMI ($kg/m^2$), mean (SD)** | 27.3 (5.2) | 27 (4.8) |
| **Endline BMI ($kg/m^2$), mean (SD)** | 27.4 (5.2) | 27.1 (4.7) |
| **Days between BMI measurements, mean (SD)** | 893.5 (601.5) | 913 (698.2) |

† Healthy diet score was adopted from the American Heart Association guidelines and was created following Rutten-Jacobs et al., 2018 paper (7, 8). The table was constructed after missing values had been filled in; No single covariate had more than 2.7% missing values. BMI: body mass index

| Supplemental Table 4. Distributions of continuous outcome variables and time difference variables, N = 10,572 | | | | | |
|---|---|---|---|---|---|
| | Days between key variable measurements | | | Distribution of BMI change outcomes standardized to three-year intervals | |
| | Two BMI measurements | Accelerometer* and baseline BMI | Follow-up BMI and accelerometer | BMI change kg/m$^2$ | Relative BMI change percentage |
| Mean | 896 | 14 | 882 | 0.03 | 0.34 |
| STD | 616 | 159 | 613 | 2.69 | 9.56 |
| Minimum | 182 | -365 | 183 | -12.21 | -44.22 |
| 25% | 441 | -90 | 462 | -1.04 | -3.91 |
| 50% | 732 | 11 | 698 | 0.06 | 0.23 |
| 75% | 1105 | 123 | 1084 | 1.27 | 4.81 |
| maximum | 3652 | 365 | 3536 | 12.09 | 45.28 |
| *The term 'accelerometer' refers to the accelerometer study date, which varied for each participant, ranging from June 2013 to December 2015. | | | | | |

| Supplemental Table 5. Associations between irregular sleep duration and risk of a standardized BMI change of at least 4.43 $kg/m^2$ standardized to three-year intervals, N = 10,572 | | | | |
|---|---|---|---|---|
| Exposure | Incident Obesity / Sample Size | RR (95% CI) Model 1† | RR (95% CI) Model 2‡ | RR (95% CI) Model 3§ |
| Sleep duration SD categories | | | | |
| ≤30 mins | 53 / 1523 | Ref. | Ref. | Ref. |
| 31-45 mins | 124 / 3138 | 1.09 (0.80, 1.48) | 1.08 (0.79, 1.48) | 1.08 (0.79, 1.47) |
| 46-60 mins | 143 / 2744 | **1.41 (1.04, 1.91)** | **1.38 (1.02, 1.87)** | **1.38 (1.02, 1.86)** |
| >60 mins | 163 / 3167 | **1.35 (1.003, 1.83)** | 1.30 (0.96, 1.76) | 1.26 (0.93, 1.71) |
| Per 1-hour increment | 483 / 10572 | 1.09 (0.95, 1.26) | 1.08 (0.93, 1.25) | 1.05 (0.90, 1.22) |
| *P*-trend* | NA | 0.2180 | 0.3003 | 0.5251 |

* Continuous sleep duration SD was included when calculating *P*-trend.
† Model 1 adjusted for age in years, sex, ethnic background, Townsend deprivation index, education, occupation/shift work, and follow-up period (days between the two BMI measurements)
‡ Model 2 adjusted for covariates in model 1 along with, smoking, physical activity, healthy diet score, alcohol consumption, dyslipidemia, hypertension, depression, and diabetes.
§ Model 3 adjusted for covariates in model 2 along with sleep apnea, insomnia, accelerometer measured sleep duration, chronotype, and daytime dozing.
Results from Poisson regression with a robust error variance model. Of note, 4.43 $kg/m^2$ is the $95^{th}$ percentile of BMI change.
SD: Standard deviation, BMI: Body mass index, RR: risk ratio, CI: confidence interval, NA: Not applicable
